# Location-specific Longitudinal Patterns of Subcortical and Lobar Cerebral Microhemorrhages in Preclinical Alzheimer Disease

**DOI:** 10.64898/2026.10.12.26365339

**Authors:** Callum Hill, Sofia Michopoulou, Mahesan Niranjan, Christopher M. Kipps

**Affiliations:** Clinical and Experimental Sciences, Faculty of Medicine, University of Southampton, Southampton, UK; School of Electronics and Computer Science, University of Southampton, Southampton, UK; Department of Medical Physics, University Hospital Southampton NHS Foundation Trust, Southampton; University Hospital Southampton NHS Foundation Trust, Southampton, UK

## Abstract

**Background and Objectives:** Cerebral microhaemorrhages influence eligibility and monitoring for anti-amyloid therapies. Emerging evidence and expert opinion suggest that subcortical microhaemorrhages may be less closely related to cerebral amyloidosis than lobar lesions and may therefore warrant separate consideration in anti-amyloid treatment eligibility criteria. Using longitudinal MRI data from cognitively unimpaired individuals with preclinical Alzheimer disease in the A4 study, we hypothesised that lobar and subcortical microhaemorrhages would show distinct location-specific patterns of progression and associated risk profiles.

**Methods:** We conducted a retrospective longitudinal cohort analysis of centrally assessed MRI data from A4 study participants with at least two evaluable MRI examinations. Baseline and follow-up were defined as the first and last evaluable scans, respectively. Participants were classified according to the anatomical distribution of baseline microhaemorrhages as lobar or subcortical, and subsequent location-specific lesion accumulation was examined. Multivariable logistic regression was used to assess whether baseline lobar and subcortical microhaemorrhage counts were independently associated with the development of additional lobar and subcortical microhaemorrhages.

**Results:** Among 1,146 participants followed for a mean of 4.55 years (SD = 1.78), 38 had subcortical-only and 157 had lobar-only microhaemorrhages at baseline. Subsequent lesions preferentially accumulated in the same anatomical compartment. Baseline subcortical microhaemorrhage count was associated with additional subcortical lesions (OR 3.60, 95% CI 2.00–6.47), but not with additional lobar lesions (OR 1.25, 95% CI 0.73–2.15). Conversely, baseline lobar microhaemorrhage count was associated with additional lobar lesions (OR 2.74, 95% CI 2.06–3.64), but not with additional subcortical lesions (OR 1.08, 95% CI 0.77–1.53). White matter hyperintensity burden was associated with subcortical progression, while APOE ε4 homozygosity and greater cerebral amyloid burden were associated with lobar progression. Systolic blood pressure was directionally positive but not associated with lobar microhaemorrhage accumulation in either analysis.

**Discussion:** Our findings support the emerging view that subcortical and lobar microhaemorrhages exhibit distinct longitudinal patterns and associated risk profiles. Longitudinal studies in patients receiving contemporary plaque-clearing therapies are needed to determine whether microhaemorrhages should be considered differently according to anatomical location in treatment eligibility and monitoring guidelines.

## Introduction

Cerebral microhaemorrhages are common MRI markers of cerebral small-vessel disease, including cerebral amyloid angiopathy. [1,2] In the context of Alzheimer’s Disease, cerebral microhaemorrhages are an important indicator of haemorrhagic risk, and microhaemorrhage counts are used to stratify patient eligibility for anti-amyloid therapies. [3]

Growing evidence suggests that cerebral microhaemorrhages may reflect different underlying vasculopathies according to their anatomical distribution. Lobar microhaemorrhages are typically associated with cerebral amyloid angiopathy and are relevant to amyloid-related imaging abnormalities during anti-amyloid treatment, whereas deep or subcortical lesions are more commonly linked to hypertensive arteriopathy. [1,4,5] These processes frequently coexist in older adults, including cognitively unimpaired individuals with elevated cerebral amyloid burden. [6] Previous studies have linked lobar microhaemorrhages with APOE ε4 and greater cerebral amyloid burden. [6, 7] Deep or subcortical microhaemorrhages have been more consistently associated with vascular risk factors such as hypertension. [7,8]

However, longitudinal evidence on whether subcortical microhaemorrhages are associated with subsequent lobar microhaemorrhages is limited. More broadly, the location-specific evolution of cerebral microhaemorrhages and the factors associated with their progression remain incompletely characterised.

Current anti-amyloid treatment eligibility criteria and ARIA-H severity classifications are based largely on total cerebral microhaemorrhage counts and do not distinguish lobar from deep or subcortical lesions despite their potentially different aetiologies. [9, 10] Expert recommendations have questioned whether microhaemorrhages in the deep grey matter and brainstem should contribute to anti-amyloid therapy eligibility counts as these lesions are considered more likely to reflect hypertensive arteriopathy than cerebral amyloidosis. [11]

The A4 study was a randomised controlled trial of solanezumab which enrolled participants with preclinical Alzheimer’s disease. [12] Solanezumab is associated with similar rates of ARIA-H to placebo and did not produce substantial cerebral amyloid clearance. [12] The trial therefore provides a large, closely monitored cohort with standardised longitudinal MRI assessments in which to examine the location-specific evolution of cerebral microhaemorrhages.

We therefore examined the longitudinal distribution of cerebral microhaemorrhages in cognitively unimpaired, amyloid-positive participants from the A4 study. We assessed whether baseline subcortical and lobar microhaemorrhage counts were differentially associated with subsequent location-specific lesion accumulation and examined the demographic, vascular, genetic, and amyloid-related factors associated with each outcome. We hypothesised that baseline lesions would preferentially predict progression in the same anatomical region and that lobar and subcortical microhaemorrhages would show distinct associated risk profiles.

## Methods

### Standard Protocol Approvals, Registrations, and Patient Consents

The A4 study (ClinicalTrials.gov: NCT02008357) was approved by the relevant institutional review boards, and all participants provided written informed consent. The reported secondary analysis was approved by the University of Southampton Faculty of Medicine Ethics Committee (ERGO:112166).

### Study design and participants

We conducted a retrospective longitudinal cohort analysis of prospectively collected MRI data from the A4 study. The A4 study enrolled cognitively normal participants with elevated cerebral amyloid burden, with MRI scans assessed centrally by study radiologists. [12] We included A4 study participants with at least two evaluable MRI assessments. The descriptive cohort included all eligible participants, whereas multivariable analyses were restricted to participants with complete outcome and covariate data. Further details of the study design can be found in the reported A4 study results. [12]

### MRI processing and baseline phenotypes

MRI findings were obtained from the reported study assessments. An MRI scan was considered evaluable when total, subcortical, and lobar microhaemorrhage counts were all available. MRI visits were ordered chronologically for each participant. The first evaluable MRI was defined as baseline and the last evaluable MRI as the latest assessment, with follow-up duration calculated as the time between these data points.

Participants were classified as having no baseline microhaemorrhage, subcortical-only microhaemorrhage, lobar-only microhaemorrhage, or both subcortical and lobar microhaemorrhages. Subcortical-only and lobar-only denoted at least one recorded lesion in the specified location and no recorded lesion in the other location.

### Statistical analysis

Baseline characteristics were described for the full eligible cohort and separately for participants with subcortical-only and lobar-only baseline microhaemorrhages. Longitudinal outcomes were summarised as numbers and proportions within each baseline phenotype for descriptive analysis. These outcomes comprised at least one additional subcortical microhaemorrhage, at least one additional lobar microhaemorrhage, at least two additional total microhaemorrhages, and no additional microhaemorrhage between baseline and the latest evaluable MRI. We compared the proportion developing at least one additional lobar microhaemorrhage between participants with subcortical-only and lobar-only baseline lesions using a two-sided Fisher’s exact test. We calculated the risk ratio and absolute risk difference with 95% confidence intervals.

The primary longitudinal endpoints were at least one additional subcortical microhaemorrhage and at least one additional lobar microhaemorrhage between the baseline and latest evaluable MRI. We fitted separate multivariable logistic regression models for each endpoint. Models included treatment assignment, age per 10 years, amyloid burden per 10 Centiloids, systolic blood pressure per 10 mm Hg, corrected WMH volume, APOE ε4 status, follow-up duration, baseline subcortical microhaemorrhage count, and baseline lobar microhaemorrhage count. Covariates were selected on the basis of established or plausible associations with cerebral microhaemorrhage burden.

We also fitted separate logistic regression models for the presence of at least one subcortical or lobar microhaemorrhage at any MRI visit. Participants were included in each model if the corresponding location-specific outcome and all model covariates were available. These models included the same covariates but excluded baseline microhaemorrhage counts as baseline observations contributed to the outcomes.

We conducted sensitivity analyses using thresholds of at least two subcortical or lobar microhaemorrhages at any evaluable visit. For the regression models, we assessed multicollinearity using variance inflation factors and pairwise correlations among continuous predictors. Statistical analysis was conducted in R version 4.5.2. This study is reported in accordance with the Strengthening the Reporting of Observational Studies in Epidemiology guidelines. [13] (Supplementary File 1)

## Data Availability

Deidentified A4 participant-level data are available to qualified researchers through the Alzheimer’s Clinical Trial Consortium A4/LEARN data portal, subject to proposal approval and applicable data-use agreements.

## Results

### Study population and baseline characteristics

Of 1,169 A4 participants with at least one dated MRI microhaemorrhage assessment, 23 had fewer than two evaluable MRI assessments and were excluded from longitudinal analyses, leaving 1,146 participants for descriptive analyses. Across the MRI source data, 6,414 dated scan rows were available, of which 6,407 had complete total, subcortical, and lobar microhaemorrhage counts. Seven scan rows were excluded because of incomplete core microhaemorrhage data. Adjusted regression analyses were restricted to complete cases for the relevant outcome and covariates; this excluded a further seven participants, leaving 1,139 participants. These participants contributed 6,348 evaluable MRI assessments to the any-visit regression analyses and 2,278 first/latest MRI assessments to the longitudinal regression analyses. (figure 1)

**Figure 1:**
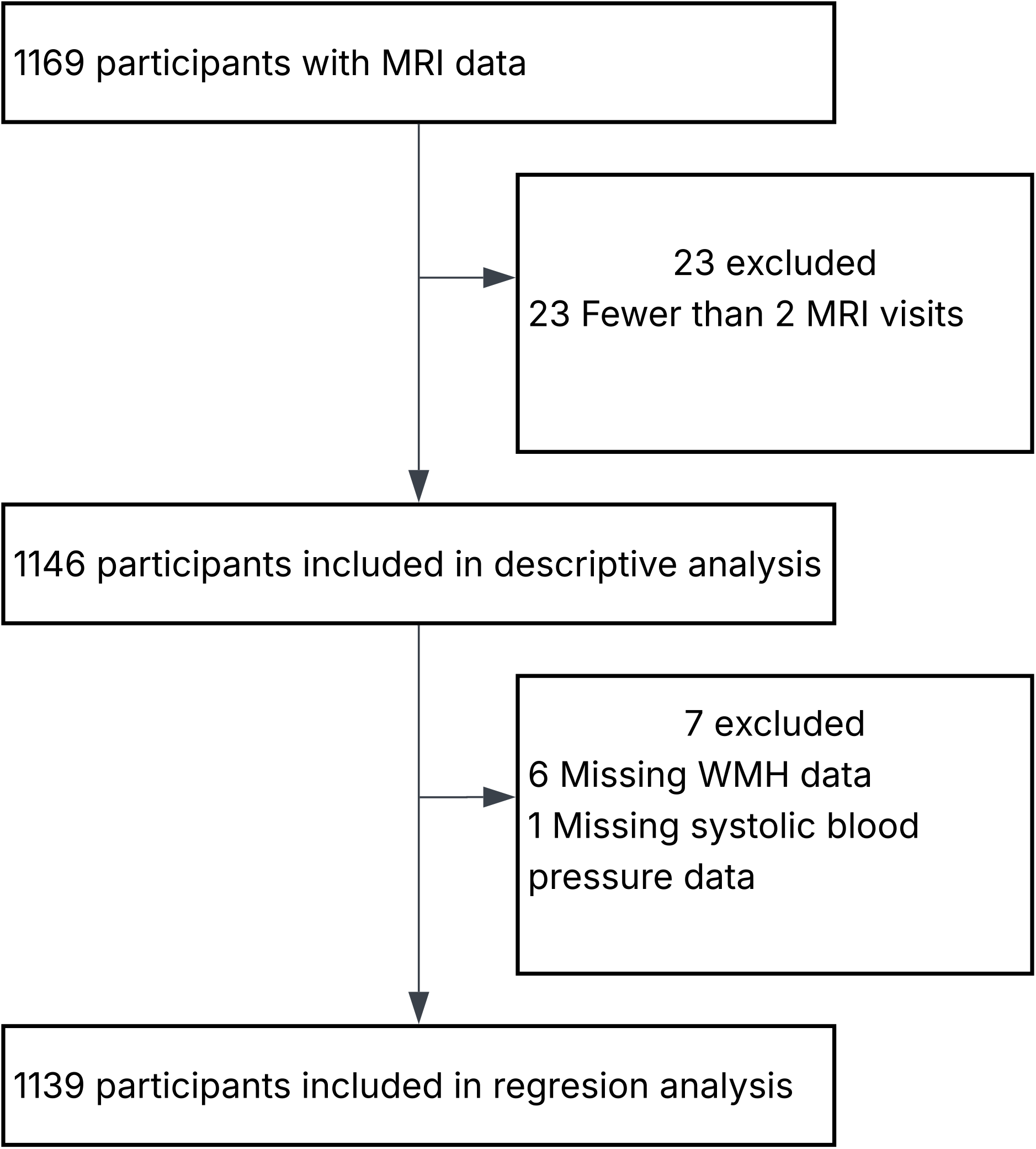
Study cohort and analytic sample processing. WMH=white matter hyperintensity.

The mean age of participants was 71.9 years (SD = 4.8), 681 (59.4%) participants were female, and the mean baseline amyloid burden was 66.1 Centiloids (SD = 32.8). At baseline, 890 (77.7%) participants had no microhaemorrhages, 181 (15.8%) had one, and 75 (6.5%) had two or more. Thirty-eight participants had subcortical-only microhaemorrhages and 157 had lobar-only microhaemorrhages. Baseline characteristics including amyloid Centiloid, white matter hyperintensities, and systolic blood pressure were broadly similar in participants with subcortical-only and lobar-only microhaemorrhages. Participants with lobar-only microhaemorrhages had a greater baseline lesion burden and a higher prevalence of APOE ε4 homozygosity. (table 1)

**Table 1:** Baseline characteristics of the analytic cohort. Data are mean (SD) or n (%). Subcortical-only denotes at least one recorded subcortical microhaemorrhage and no recorded lobar microhaemorrhage; lobar-only denotes at least one recorded lobar microhaemorrhage and no recorded subcortical microhaemorrhage. APOE=apolipoprotein E.

| Characteristic | All participants (N=1,146) | Subcortical-only baseline microhaemorrhage (N=38) | Lobar-only baseline microhaemorrhage (N=157) |
| --- | --- | --- | --- |
| Age, years | 71.9 (4.8) | 73.8 (5.4) | 72.5 (4.9) |
| Amyloid burden, Centiloids | 66.1 (32.8) | 70.5 (41.3) | 68.8 (32.6) |
| White matter hyperintensity (WMH), cm<sup>3</sup> | 5.0 (6.3) | 6.9 (6.7) | 5.7 (7.1) |
| Sex, n (%) |  |  |  |
| Female | 681 (59.4%) | 21 (55.3%) | 76 (48.4%) |
| Male | 465 (40.6%) | 17 (44.7%) | 81 (51.6%) |
| Race, n (%) |  |  |  |
| White | 1080 (94.2%) | 36 (94.7%) | 143 (91.1%) |
| Black | 27 (2.4%) | 0 (0.0%) | 7 (4.5%) |
| Asian | 23 (2.0%) | 2 (5.3%) | 5 (3.2%) |
| Other | 16 (1.4%) | 0 (0.0%) | 2 (1.3%) |
| **APOE ε4 status, n (%)** |  |  |  |
| Non-carrier | 472 (41.2%) | 23 (60.5%) | 52 (33.1%) |
| Heterozygote | 580 (50.6%) | 12 (31.6%) | 82 (52.2%) |
| Homozygote | 94 (8.2%) | 3 (7.9%) | 23 (14.6%) |
| Systolic blood pressure, mmHg | 136.6 (16.1) | 137.2 (16.8) | 137.3 (16.2) |
| **Baseline microhaemorrhage count, n (%)** |  |  |  |
| 0 | 890 (77.7%) | 0 (0.0%) | 0 (0.0%) |
| 1 | 181 (15.8%) | 34 (89.5%) | 102 (65.0%) |
| 2 or more | 75 (6.5%) | 4 (10.5%) | 55 (35.0%) |
| **Baseline superficial siderosis, n (%)** |  |  |  |
| 0 | 1131 (98.7%) | 37 (97.4%) | 151 (96.2%) |
| 1 or more | 15 (1.3%) | 1 (2.6%) | 6 (3.8%) |
| **Treatment assignment, n (%)** |  |  |  |
| Solanezumab | 563 (49.1%) | 17 (44.7%) | 64 (40.8%) |
| Placebo | 583 (50.9%) | 21 (55.3%) | 93 (59.2%) |

### Longitudinal outcomes by baseline phenotype

Longitudinal patterns of microhaemorrhage accumulation differed according to baseline phenotype (table 2). Mean follow-up time from baseline to final MRI was 4.55 years (SD= 1.78).

**Table 2:** Additional cerebral microhaemorrhages according to baseline phenotype. Data are n (%). Additional microhaemorrhages were defined by the change from baseline to the final MRI.

| Baseline phenotype | N | ≥1 additional subcortical microhaemorrhage, n (%) | ≥1 additional lobar microhaemorrhage, n (%) | ≥2 additional total microhaemorrhages, n (%) | No additional microhaemorrhages, n (%) |
| --- | --- | --- | --- | --- | --- |
| No baseline microhaemorrhage | 8<br>9<br>0 | 33 (3.7%) | 125 (14.0%) | 89 (10.0%) | 666 (74.8%) |
| Subcortical-only microhaemorrhage | 3<br>8 | 6 (15.8%) | 9 (23.7%) | 9 (23.7%) | 24 (63.2%) |
| Lobar-only microhaemorrhage | 1<br>5<br>7 | 9 (5.7%) | 68 (43.3%) | 52 (33.1%) | 69 (43.9%) |
| Subcortical and lobar microhaemorrhage | 1<br>4 | 4 (28.6%) | 3 (21.4%) | 4 (28.6%) | 7 (50.0%) |

Among participants without baseline microhaemorrhages, 3.7% developed at least one additional subcortical microhaemorrhage and 14.0% developed at least one additional lobar microhaemorrhage. Subsequent subcortical microhaemorrhage occurred in 15.8% of participants with subcortical-only baseline lesions and 5.7% of those with lobar-only lesions. By contrast, subsequent lobar microhaemorrhage occurred in 23.7% of the subcortical-only group and 43.3% of the lobar-only group.

We compared subsequent lobar microhaemorrhage risk between participants with subcortical-only and lobar-only baseline lesions using a two-sided unadjusted Fisher’s exact test. Subsequent lobar microhaemorrhage occurred in 9 of 38 participants with subcortical-only lesions and 68 of 157 with lobar-only lesions, corresponding to a risk ratio of 0.55 (95% CI 0.30–0.99) and a risk difference of −19.6 percentage points (95% CI −35.2 to −4.0; p=0.028).

### Adjusted associations with location-specific progression

We examined adjusted associations between covariates and the presence of at least one additional subcortical or lobar microhaemorrhage in separate multivariable models. Among 1139 participants in the adjusted analyses, 56 developed at least one additional subcortical microhaemorrhage and 218 developed at least one additional lobar microhaemorrhage. For subcortical progression (figure 2A), baseline subcortical microhaemorrhage count was strongly associated with additional subcortical microhaemorrhage (OR: 3.60, 95% CI: 2.00-6.47), whereas baseline lobar count was not (OR: 1.08, 95% CI: 0.77-1.53). Longer follow-up was associated with further subcortical microhaemorrhages (1.46, 1.15-1.86), and greater WMH burden showed a significant positive association (1.03, 1.00-1.07). Treatment assignment, age, amyloid burden, systolic blood pressure, and APOE ε4 status were not clearly associated with additional subcortical microhaemorrhage.

**Figure 2.**
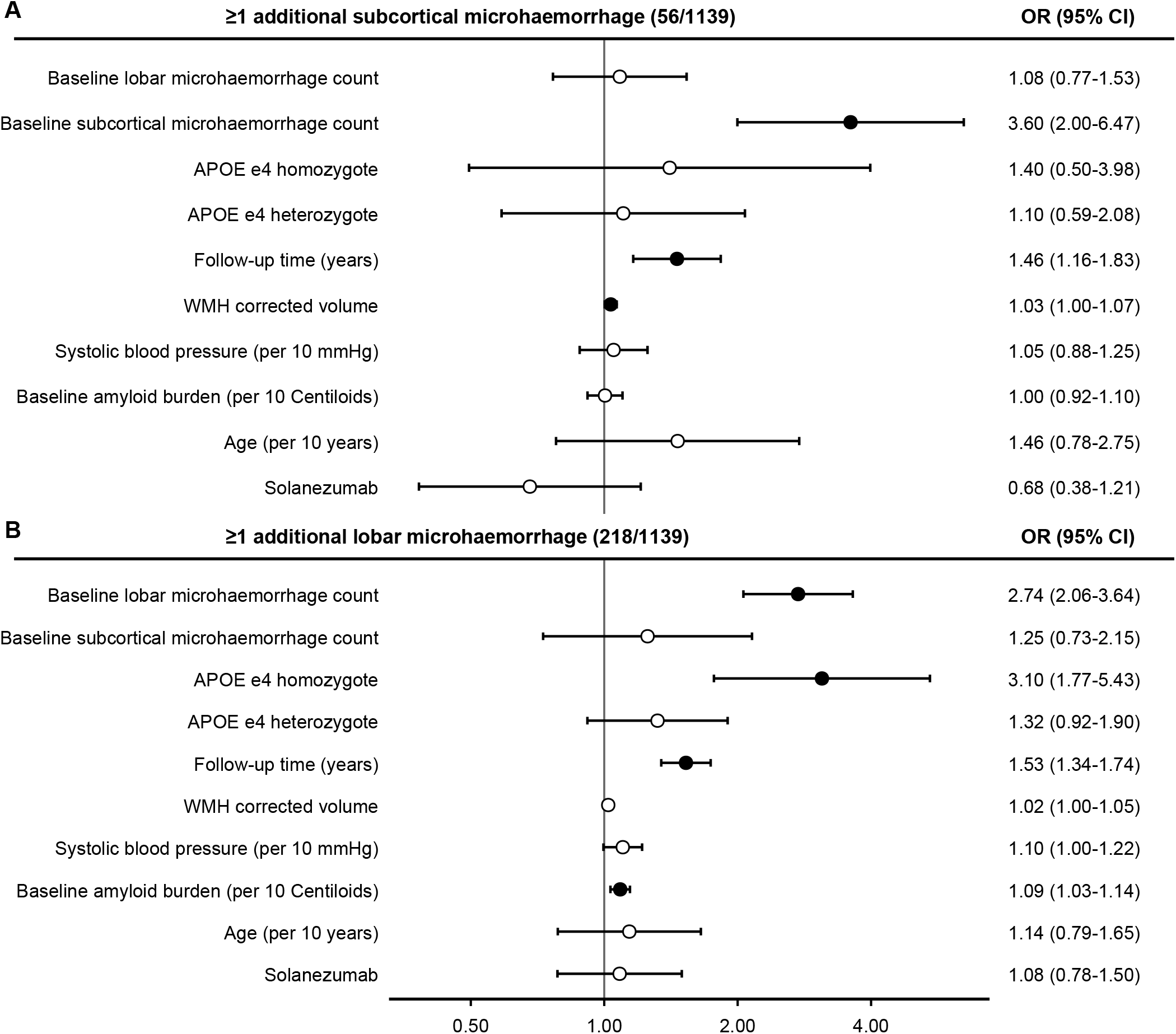
Adjusted longitudinal associations with at least 1 additional subcortical or lobar cerebral microhaemorrhage from baseline to final MRI. Forest plots show adjusted odds ratios and 95% CIs from multivariable logistic regression models for additional subcortical microhaemorrhage (A) and additional lobar microhaemorrhage (B). Filled circles indicate statistically significant results. WMH=white matter hyperintensity (cm^3^).

For lobar progression (figure 2B), baseline lobar microhaemorrhage count was strongly associated with additional lobar microhaemorrhage (OR 2.74, 95% CI 2.06-3.64), whereas baseline subcortical count was not (OR: 1.25, 95% CI: 0.73-2.15). APOE ε4 homozygosity (3.10, 1.77-5.43), greater amyloid burden per 10 Centiloids (1.09, 1.03-1.15), and longer follow-up (1.53, 1.34-1.74) were also associated with lobar progression. The estimates for APOE ε4 heterozygosity, treatment assignment, and age were non-significant.

### Any-visit microhaemorrhage outcomes and sensitivity analysis

In secondary analyses, we analysed factors associated with the presence of at least one subcortical or lobar microhaemorrhage at any MRI visit (figure 3). Among 1139 participants, 98 had at least one subcortical microhaemorrhage and 319 had at least one lobar microhaemorrhage at any visit. For subcortical microhaemorrhage (figure 3A), older age (OR 1.65 per 10 years, 95% CI 1.04-2.60) and greater WMH burden (OR 1.04, 95% CI 1.01-1.06) were associated with an increased risk of subcortical microhaemorrhages.

**Figure 3.**
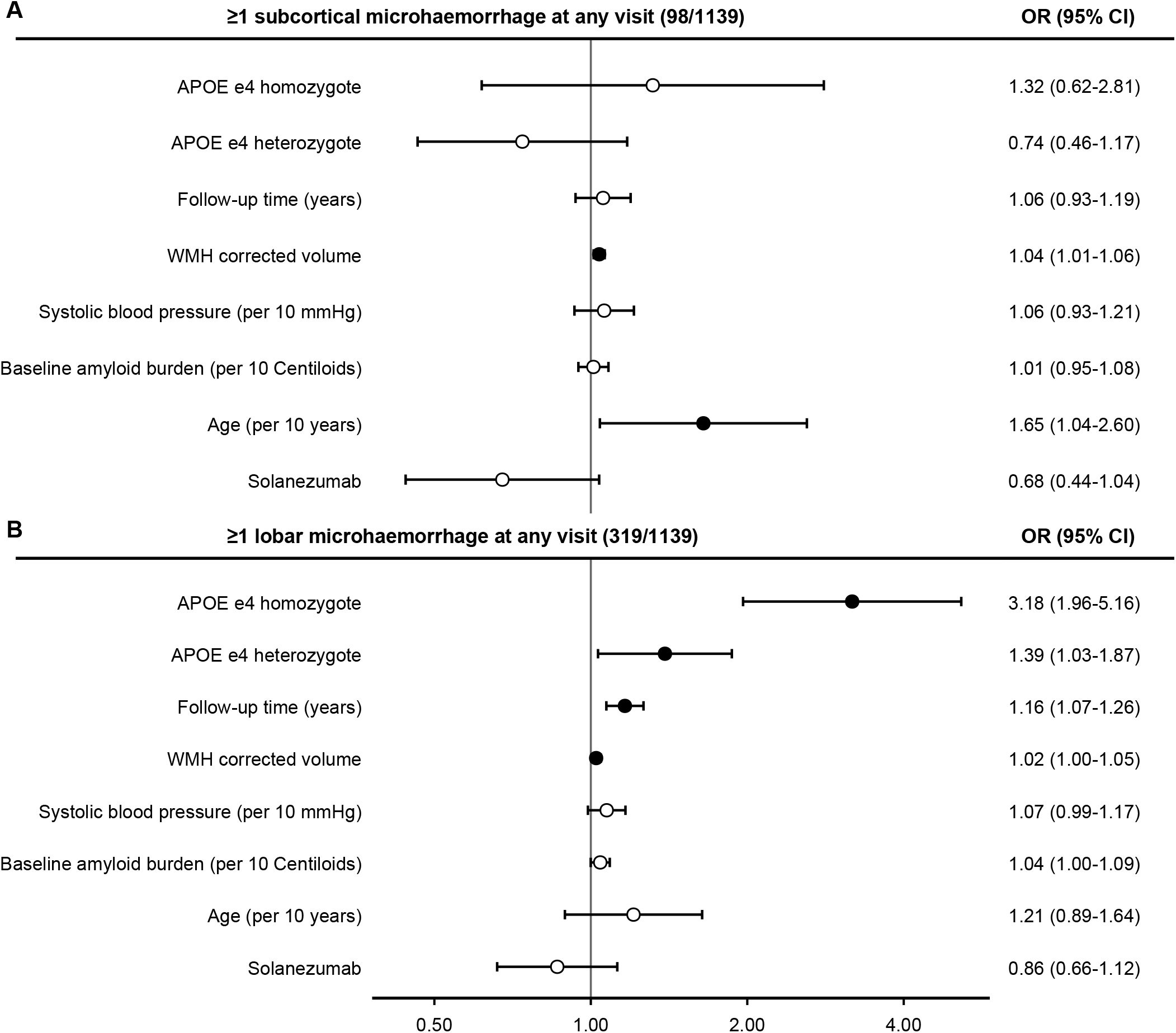
Adjusted associations with subcortical or lobar cerebral microhaemorrhage at any MRI visit. Forest plots show adjusted odds ratios and 95% CIs from multivariable logistic regression models for at least 1 subcortical microhaemorrhage at any visit (A) and at least 1 lobar microhaemorrhage at any visit (B). Models included all variables shown. Filled circles indicate statistically significant associations. WMH=white matter hyperintensity (cm^3^).

For lobar microhaemorrhage (figure 3B), APOE ε4 homozygosity (OR 3.18, 95% CI 1.96-5.16) and heterozygosity (OR 1.39, 95% CI 1.03-1.87) were associated with higher odds of having at least one lobar microhaemorrhage at any MRI visit. Longer follow-up was also associated with higher odds of microhaemorrhage events (OR 1.16 per year, 95% CI 1.07-1.26), with smaller positive associations for WMH burden (OR 1.02, 95% CI 1.00-1.05) and amyloid burden (OR 1.04 per 10 Centiloids, 95% CI 1.00-1.09).

We conducted sensitivity analyses using a higher threshold of at least two microhaemorrhages at any MRI visit (Supplementary File 2). Estimates in the subcortical models were less precise due to the small number of events, and longitudinal multivariable analysis was not possible using higher count thresholds due to the small number of events (n=9). Compared with the primary any-visit analyses, endpoint sensitivity analysis of at least two microhaemorrhages showed a broadly similar pattern but with greater imprecision. For subcortical microhaemorrhages, WMH burden remained associated with subcortical microhaemorrhage, while other covariates were non-significant, likely due to the small number of events (n=20). For lobar microhaemorrhages, APOE ε4 homozygosity showed a stronger association at the higher threshold, while the association with APOE ε4 heterozygosity was no longer statistically significant. Longer follow-up and WMH burden remained associated with lobar microhaemorrhage burden, whereas the association with amyloid burden was weaker.

## Discussion

In this longitudinal analysis of cognitively unimpaired, amyloid-positive participants from the A4 study, cerebral microhaemorrhages showed a clear pattern of preferential same-location progression. The location-specific associations were substantial, particularly for baseline subcortical microhaemorrhages and subsequent subcortical lesion accumulation. The associated risk profiles also differed: lobar progression was associated with APOE ε4 homozygosity and greater cerebral amyloid burden, whereas subcortical progression was associated with greater WMH burden. Together, these findings suggest that lobar and subcortical microhaemorrhages may reflect partially distinct underlying small-vessel disease processes.

The descriptive analyses were consistent with a similar pattern of location-specific progression. Subsequent subcortical microhaemorrhages occurred in 15.8% of participants with subcortical-only baseline lesions, compared with 5.7% of those with lobar-only lesions. Conversely, subsequent lobar microhaemorrhages occurred in 43.3% of participants with lobar-only baseline lesions, compared with 23.7% of those with subcortical-only lesions. In the unadjusted Fisher exact comparison, participants with subcortical-only lesions had a lower risk of subsequent lobar microhaemorrhage than those with lobar-only lesions. However, this result should be interpreted in the context of a more limited sample size in the subcortical-only phenotype and the differing baseline characteristics of each group, particularly with regard to APOE ε4 status and baseline lesion burden.

The pattern of lobar microhaemorrhage accumulation was broadly consistent with established risk factors for haemorrhagic vascular injury in the context of cerebral amyloidosis. APOE ε4 homozygosity was strongly associated with lobar microhaemorrhages in both the longitudinal and any-visit analyses. Higher cerebral amyloid burden was also associated with lobar progression, although the estimate was weaker in the any-visit analysis. Together, this pattern is consistent with previous work linking APOE ε4, cerebral amyloidosis, and lobar haemorrhagic lesions, and supports the interpretation that established lobar lesion burden identifies ongoing amyloid-related vascular vulnerability. [6, 14, 15] WMH volume showed a smaller positive association, reaching significance in the any-visit analysis, suggesting that lobar microhaemorrhages may also arise within a broader background of cerebral small-vessel injury. Systolic blood pressure was directionally positive but not independently associated with lobar outcomes in either analysis.

The subcortical regression models showed a different pattern, more consistent with non-amyloid small-vessel injury. Greater WMH volume was associated with subcortical microhaemorrhages in both the longitudinal and any-visit analyses, supporting the occurrence of these lesions in the context of cerebral small-vessel disease. Older age was associated with the any-visit outcome but not clearly with longitudinal progression, potentially reflecting its nature as an indicator of cumulative lesion prevalence, although this may simply reflect the smaller sample size in the longitudinal analysis. Amyloid burden and APOE ε4 status were not independently associated with subcortical outcomes, supporting a distinction from the amyloid-related profile observed for lobar lesions. Systolic blood pressure was not independently associated with subcortical outcomes in either model. This null finding should be interpreted in the context of the smaller number of subcortical events, limiting power to detect modest associations. It should be considered in interpreting this result that a single baseline clinic reading in an amyloid-selected trial cohort, many likely treated for hypertension, is not a robust measure of lifetime hypertensive exposure. In contrast, the consistent association with WMH burden suggests that cumulative cerebral small-vessel injury may be more informative than contemporaneous blood pressure in this cohort.

Several mechanisms may account for the observed location-specific patterns. Greater lobar microhaemorrhage burden has been linked pathologically to cortical, leptomeningeal, and particularly capillary CAA, supporting an amyloid-related mechanism for lobar progression. [2,16] By contrast, deep microhaemorrhages are more commonly associated with arteriolosclerotic injury in perforating vessels. [17-19] However, our findings do not provide evidence for wholly separate lobar and subcortical disease processes: lesion location is an indirect marker, mixed vasculopathies are common, and the association of WMH burden with both outcomes may reflect shared downstream small-vessel damage. [20]

Our findings support the emerging view that subcortical microhaemorrhages may represent a distinct non-amyloid-related small vessel disease, and as such their inclusion in microhaemorrhage counts in determining anti-amyloid eligibility may warrant reconsideration. [11] However, our results should be interpreted cautiously; our findings add to the evidence that subcortical microhaemorrhages may warrant separate consideration within existing eligibility thresholds, but do not establish that current eligibility criteria should be changed. As solanezumab demonstrated relatively limited amyloid clearance, our results represent progression trajectories in the absence of effective anti-amyloid clearing therapies. [12] Further evaluation of anatomical lesion stratification in cohorts receiving contemporary plaque-clearing therapies is needed before evidence-based recommendations can be made regarding the treatment implications of subcortical lesions. This issue is likely to become increasingly important as novel and emerging anti-amyloid therapies extend treatment eligibility to broader and more clinically heterogeneous patient populations. [11] We suggest that further studies of regional microhaemorrhage classification may provide evidence to support changes in treatment eligibility guidelines and location-specific monitoring requirements for incident microhaemorrhages.

Our results extend previous cross-sectional and longitudinal studies. The Rotterdam Scan Study similarly found deep or infratentorial microhaemorrhages were preferentially associated with cardiovascular risk factors and white matter lesions. [17] The Mayo Clinic Study of Aging provided evidence for the association of baseline cerebral microhaemorrhages with future lesions in a cohort not selected for cerebral amyloidosis. [14] However, longitudinal evidence for the comparative progression of subcortical and lobar microhaemorrhages in the context of Alzheimer’s disease or cerebral amyloidosis remains limited in the current literature. Gay et al. 2025 found a higher cross-sectional prevalence of both lobar and deep microhaemorrhages in amyloid-positive A4 participants than in amyloid-negative LEARN participants, although APOE ε4 homozygosity was associated only with lobar lesions. [4] Yates et al. 2014 in the Australian Imaging, Biomarkers and Lifestyle Study of Ageing demonstrated amyloid-associated longitudinal accumulation of lobar microhaemorrhages in an Alzheimer-spectrum cohort, but the small number of deep or infratentorial events precluded a direct longitudinal comparison of lobar and deep lesion progression. [15]

Strengths of this study include the large, well-monitored cohort of individuals with elevated cerebral amyloid with a mean follow-up period of over 4 years in which to assess the progression of haemorrhagic lesions. Centralised, standardised MRI interpretation enhanced the reliability and consistency of lesion assessment, while protocolised covariate collection resulted in minimal missing data and supported rigorous longitudinal analysis. The complementary longitudinal and any-visit analyses also distinguished factors associated with subsequent accumulation from those associated with cumulative lesion presence.

However, several limitations should be considered. This was a secondary observational analysis and cannot establish causal relationships between the measured covariates and microhaemorrhage progression. The number of subcortical microhaemorrhages was relatively small, limiting the ability to detect modest associations. Due to the small number of participants with subcortical microhaemorrhage accumulation, a sensitivity analysis was not possible for alternative higher threshold endpoints in the longitudinal analysis. Longitudinal change was defined between the first and last evaluable MRI and therefore did not capture the timing or clustering of lesion development between intermediate scans. As pathological confirmation of cerebral amyloid angiopathy or hypertensive arteriopathy was unavailable, the observed haemorrhagic lesion patterns cannot be attributed definitively to a specific underlying vasculopathy or mechanism. Finally, the selected, predominantly white, cognitively unimpaired A4 study population and the limited amyloid-clearing effect of solanezumab restrict generalisability to symptomatic Alzheimer’s disease and to patients receiving contemporary amyloid targeting therapies.

## Conclusion

In cognitively unimpaired, amyloid-positive participants receiving treatment with solanezumab or placebo, baseline microhaemorrhage location was associated with the anatomical distribution of subsequent lesions. Lobar outcomes were more closely associated with APOE ε4 homozygosity and cerebral amyloid burden, whereas subcortical outcomes were more closely associated with WMH burden. Our findings support preferential same-location progression and distinct associated risk patterns, while the association of WMH burden with both lesion types suggests that the underlying vascular processes may partially overlap. Longitudinal studies in cohorts receiving contemporary amyloid-clearing therapies should evaluate whether baseline lobar and subcortical microhaemorrhages differ in their associations with microhaemorrhage accumulation and ARIA more broadly. External validation across antibodies and clinically representative populations is needed before anatomical distribution of haemorrhagic lesions can be reliably incorporated into treatment eligibility recommendations and clinical guidelines.

## Acknowledgements

The A4 Study was a secondary prevention trial in preclinical Alzheimer’s disease, aiming to slow cognitive decline associated with brain amyloid accumulation in clinically normal older individuals. The A4 Study was funded by a public-private-philanthropic partnership, including funding from the National Institutes of Health-National Institute on Aging, Eli Lilly and Company, Alzheimer’s Association, Accelerating Medicines Partnership, GHR Foundation, an anonymous foundation, and additional private donors, with in-kind support from Avid Radiopharmaceuticals, Cogstate, Albert Einstein College of Medicine and the Foundation for Neurologic Diseases. The companion observational Longitudinal Evaluation of Amyloid Risk and Neurodegeneration (LEARN) Study was funded by the Alzheimer’s Association and GHR Foundation. The A4 and LEARN Studies were led by Dr. Reisa Sperling at Brigham and Women’s Hospital, Harvard Medical School, and Dr. Paul Aisen at the Alzheimer’s Therapeutic Research Institute (ATRI) at the University of Southern California. The A4 and LEARN Studies were coordinated by ATRI at the University of Southern California, and the data are made available under the auspices of Alzheimer’s Clinical Trial Consortium through the Global Research & Imaging Platform (GRIP). The complete A4 Study Team list is available at: https://www.actcinfo.org/a4-study-team-lists/. We would like to acknowledge the dedication of the study participants and their study partners who made the A4 and LEARN Studies possible.

ChatGPT (OpenAI, GPT 5.6-sol) was used to assist with proofreading of the manuscript. All suggested changes were reviewed and approved by the authors, who take full responsibility for the accuracy, originality, and final content of the manuscript.

## Sources of Funding

S.M. is funded by the National Institute for Health and Care Research (NIHR306220). The views expressed in this publication are those of the authors and not necessarily those of the funding body. The funders had no role in the design, analysis, interpretation, or reporting of the present secondary analysis.

## Conflicts of interest statement

CMK delivers industry-sponsored trials, provides consultancy and has accepted hospitality from Biogen, Eisai, Lilly, Novartis and Roche. All other authors declare no conflicts of interest.

## Notes

### Author Declarations

The Ethics Committee of the Faculty of Medicine, University of Southampton, gave ethical approval for this work (ERGO: 112166).

## References

[1] Greenberg SM, Vernooij MW, Cordonnier C, et al. Cerebral microbleeds: a guide to detection and interpretation. The Lancet Neurology. 2009;8(2):165–174. doi:10.1016/S1474-4422(09)70013-4

[2] Graff-Radford J, Lesnick TG, Mielke MM, et al. Cerebral Amyloid Angiopathy Burden and Cerebral Microbleeds: Pathological Evidence for Distinct Phenotypes. Ikram MA, ed. JAD. 2021;81(1):113–122. doi:10.3233/JAD-201536

[3] Cogswell PM, Andrews TJ, Barakos JA, et al. Alzheimer Disease Anti-Amyloid Immunotherapies: Imaging Recommendations and Practice Considerations for Monitoring of Amyloid-Related Imaging Abnormalities. AJNR Am J Neuroradiol. 2025;46(1):24–32. doi:10.3174/ajnr.A8469

[4] Gay MD, Baldaranov D, Donohue MC, et al. Relationship between use of anti_platelet agents, oral anti_coagulants, and Aβ burden with cerebral microhemorrhages in cognitively asymptomatic adults. Alzheimer’s & Dementia. 2025;21(5):e70167. doi:10.1002/alz.70167

[5] Agarwal A, Gupta V, Brahmbhatt P, et al. Amyloid-related Imaging Abnormalities in Alzheimer Disease Treated with Anti–Amyloid-β Therapy. RadioGraphics. 2023;43(9):e230009. doi:10.1148/rg.230009

[6] Graff-Radford J, Botha H, Rabinstein AA, et al. Cerebral microbleeds: Prevalence and relationship to amyloid burden. Neurology. 2019;92(3). doi:10.1212/WNL.0000000000006780

[7] Jensen PN, Rashid T, Ware JB, et al. Association of brain microbleeds with risk factors, cognition, and MRI markers in MESA. Alzheimer’s & Dementia. 2023;19(9):4139–4149. doi:10.1002/alz.13346

[8] Oomens JE, van Gils V, Vos SJB, et al. Cerebral Microbleeds and Amyloid Pathology Estimates From the Amyloid Biomarker Study. JAMA Netw Open. 2025;8(1):e2455571. doi:10.1001/jamanetworkopen.2024.55571

[9] Cummings J, Apostolova L, Rabinovici GD, et al. Lecanemab: Appropriate Use Recommendations. J Prev Alzheimers Dis. 2023;10(3):362–377. doi:10.14283/jpad.2023.30

[10] Rabinovici GD, Selkoe DJ, Schindler SE, et al. Donanemab: Appropriate use recommendations. J Prev Alzheimers Dis. 2025;12(5):100150. doi:10.1016/j.tjpad.2025.100150

[11] Cogswell PM, Barakos JA, Barkhof F, et al. Amyloid-Related Imaging Abnormalities with Emerging Alzheimer Disease Therapeutics: Detection and Reporting Recommendations for Clinical Practice. AJNR Am J Neuroradiol. 2022;43(9):E19–E35. doi:10.3174/ajnr.A7586

[12] Sperling RA, Donohue MC, Raman R, et al. Trial of Solanezumab in Preclinical Alzheimer’s Disease. N Engl J Med. 2023;389(12):1096–1107. doi:10.1056/NEJMoa2305032

[13] Elm EV, Altman DG, Egger M, Pocock SJ, Gøtzsche PC, Vandenbroucke JP. Strengthening the reporting of observational studies in epidemiology (STROBE) statement: guidelines for reporting observational studies. BMJ. 2007;335(7624):806–808. doi:10.1136/bmj.39335.541782.AD

[14] Graff-Radford J, Lesnick T, Rabinstein AA, et al. Cerebral microbleed incidence, relationship to amyloid burden: The Mayo Clinic Study of Aging. Neurology. 2020;94(2):e190–e199. doi:10.1212/WNL.0000000000008735

[15] Yates PA, Desmond PM, Phal PM, et al. Incidence of cerebral microbleeds in preclinical Alzheimer disease. Neurology. 2014;82(14):1266–1273. doi:10.1212/WNL.0000000000000285

[16] Attems J, Jellinger K, Thal DR, Van Nostrand W. Review: sporadic cerebral amyloid angiopathy. Neuropathol Appl Neurobiol. 2011;37(1):75–93. doi:10.1111/j.1365-2990.2010.01137.x

[17] Poels MMF, Vernooij MW, Ikram MA, et al. Prevalence and Risk Factors of Cerebral Microbleeds: An Update of the Rotterdam Scan Study. Stroke. 2010;41(10_suppl_1). doi:10.1161/STROKEAHA.110.595181

[18] Shimoyama T, Iguchi Y, Kimura K, et al. Stroke patients with cerebral microbleeds on MRI scans have arteriolosclerosis as well as systemic atherosclerosis. Hypertens Res. 2012;35(10):975–979. doi:10.1038/hr.2012.84

[19] Vernooij MW, van der Lugt A, Ikram MA, et al. Prevalence and risk factors of cerebral microbleeds: the Rotterdam Scan Study. Neurology. 2008;70(14):1208–1214. doi:10.1212/01.wnl.0000307750.41970.d9

[20] Guidoux C, Hauw JJ, Klein IF, et al. Amyloid Angiopathy in Brain Hemorrhage: A Postmortem Neuropathological-Magnetic Resonance Imaging Study. Cerebrovasc Dis. 2018;45(3-4):124–131. doi:10.1159/000486554

