## Supplementary File 1 for "Location-specific Longitudinal Patterns of Subcortical and Lobar Cerebral Microhemorrhages in Preclinical Alzheimer Disease"

STROBE Statement—checklist of items that should be included in reports of observational studies

|  | Item No. | Recommendation | Page  No. | Relevant text from manuscript |
| --- | --- | --- | --- | --- |
| **Title and abstract** | 1 | (*a*) Indicate the study’s design with a commonly used term in the title or the abstract | 1 | We conducted a retrospective longitudinal cohort analysis of centrally assessed MRI data from A4 study participants with at least two evaluable MRI examinations. |
|  |  | (*b*) Provide in the abstract an informative and balanced summary of what was done and what was found | 1 | Abstract: methods and results |
| Introduction | | | |  |
| Background/rationale | 2 | Explain the scientific background and rationale for the investigation being reported | 2 | Cerebral microhaemorrhages are common MRI markers of cerebral small-vessel disease, including cerebral amyloid angiopathy. |
| Objectives | 3 | State specific objectives, including any prespecified hypotheses | 2 | We hypothesised that baseline lesions would preferentially predict progression in the same anatomical region and that lobar and subcortical microhaemorrhages would show distinct associated risk profiles. |
| Methods | | | |  |
| Study design | 4 | Present key elements of study design early in the paper | 3 We conducted a retrospective longitudinal cohort analysis of prospectively collected MRI data from the A4 study. The A4 study enrolled cognitively normal participants with elevated cerebral amyloid burden, with MRI scans assessed centrally by study radiologists. [12] We included A4 study participants with at least two evaluable MRI assessments. The descriptive cohort included all eligible participants, whereas multivariable analyses were restricted to participants with complete outcome and covariate data. |  |
| Setting | 5 | Describe the setting, locations, and relevant dates, including periods of recruitment, exposure, follow-up, and data collection | 3 We conducted a retrospective longitudinal cohort analysis of prospectively collected MRI data from the A4 study. The A4 study enrolled cognitively normal participants with elevated cerebral amyloid burden, with MRI scans assessed centrally by study radiologists. [12] We included A4 study participants with at least two evaluable MRI assessments. The descriptive cohort included all eligible participants, whereas multivariable analyses were restricted to participants with complete outcome and covariate data. Further details of the study design can be found in the reported A4 study results. [12] |  |
| Participants | 6 | (*a*) *Cohort study*—Give the eligibility criteria, and the sources and methods of selection of participants. Describe methods of follow-up  *Case-control study*—Give the eligibility criteria, and the sources and methods of case ascertainment and control selection. Give the rationale for the choice of cases and controls  *Cross-sectional study*—Give the eligibility criteria, and the sources and methods of selection of participants | 3 Further details of the study design can be found in the reported A4 study results. [12] MRI findings were obtained from the reported study assessments. An MRI scan was considered evaluable when total, subcortical, and lobar microhaemorrhage counts were all available. MRI visits were ordered chronologically for each participant. The first evaluable MRI was defined as baseline and the last evaluable MRI as the latest assessment, with follow-up duration calculated as the time between these data points.  Participants were classified as having no baseline microhaemorrhage, subcortical-only microhaemorrhage, lobar-only microhaemorrhage, or both subcortical and lobar microhaemorrhages. Subcortical-only and lobar-only denoted at least one recorded lesion in the specified location and no recorded lesion in the other location. |  |
|  |  | (*b*) *Cohort study*—For matched studies, give matching criteria and number of exposed and unexposed  *Case-control study*—For matched studies, give matching criteria and the number of controls per case | N/A |  |
| Variables | 7 | Clearly define all outcomes, exposures, predictors, potential confounders, and effect modifiers. Give diagnostic criteria, if applicable | 3 The primary longitudinal endpoints were at least one additional subcortical microhaemorrhage and at least one additional lobar microhaemorrhage between the baseline and latest evaluable MRI. |  |
| Data sources/ measurement | 8* | For each variable of interest, give sources of data and details of methods of assessment (measurement). Describe comparability of assessment methods if there is more than one group | *4* MRI findings were obtained from the reported study assessments. An MRI scan was considered evaluable when total, subcortical, and lobar microhaemorrhage counts were all available. |  |
| Bias | 9 | Describe any efforts to address potential sources of bias 4 | The A4 study enrolled cognitively normal participants with elevated cerebral amyloid burden, with MRI scans assessed centrally by study radiologists. |  |
| Study size | 10 | Explain how the study size was arrived at 4 | No a priori sample-size calculation; all eligible participants were included |  |

Continued on next page

| Quantitative variables | 11 | Explain how quantitative variables were handled in the analyses. If applicable, describe which groupings were chosen and why |  | 3, 4 Models included treatment assignment, age per 10 years, amyloid burden per 10 Centiloids, systolic blood pressure per 10 mm Hg, corrected WMH volume, APOE ε4 status, follow-up duration, baseline subcortical microhaemorrhage count, and baseline lobar microhaemorrhage count |
| --- | --- | --- | --- | --- |
| Statistical methods | 12 | (*a*) Describe all statistical methods, including those used to control for confounding | 3,4 | We fitted separate multivariable logistic regression models for each endpoint. Covariates were selected on the basis of established or plausible associations with cerebral microhaemorrhage burden. |
|  |  | (*b*) Describe any methods used to examine subgroups and interactions No formal subgroup interaction analyses were performed. |  |  |
|  |  | (*c*) Explain how missing data were addressed | 3 | The descriptive cohort included all eligible participants, whereas multivariable analyses were restricted to participants with complete outcome and covariate data. |
|  |  | (*d*) *Cohort study*—If applicable, explain how loss to follow-up was addressed  *Case-control study*—If applicable, explain how matching of cases and controls was addressed  *Cross-sectional study*—If applicable, describe analytical methods taking account of sampling strategy | 3 | The first evaluable MRI was defined as baseline and the last evaluable MRI as the latest assessment, with follow-up duration calculated as the time between these data points. |
|  |  | (*e*) Describe any sensitivity analyses | 4 | We conducted sensitivity analyses using thresholds of at least two subcortical or lobar microhaemorrhages at any evaluable visit. |
| Results | | | | |
| Participants | 13* | (a) Report numbers of individuals at each stage of study—eg numbers potentially eligible, examined for eligibility, confirmed eligible, included in the study, completing follow-up, and analysed | 4 Of 1,169 A4 participants with at least one dated MRI microhaemorrhage assessment, 23 had fewer than two evaluable MRI assessments and were excluded from longitudinal analyses, leaving 1,146 participants for descriptive analyses. |  |
|  |  | (b) Give reasons for non-participation at each stage | 4  Seven scan rows were excluded because of incomplete core microhaemorrhage data. |  |
|  |  | (c) Consider use of a flow diagram | 5 |  |
| Descriptive data | 14* | (a) Give characteristics of study participants (eg demographic, clinical, social) and information on exposures and potential confounders | 5,6 Table 1 |  |
|  |  | (b) Indicate number of participants with missing data for each variable of interest | 5  Figure 1 |  |
|  |  | (c) *Cohort study*—Summarise follow-up time (eg, average and total amount) | 5 |  |
| Outcome data | 15* | *Cohort study*—Report numbers of outcome events or summary measures over time | 4,5 |  |
|  |  | *Case-control study—*Report numbers in each exposure category, or summary measures of exposure |  |  |
|  |  | *Cross-sectional study—*Report numbers of outcome events or summary measures |  |  |
| Main results | 16 | (*a*) Give unadjusted estimates and, if applicable, confounder-adjusted estimates and their precision (eg, 95% confidence interval). Make clear which confounders were adjusted for and why they were included | 8,9 |  |
|  |  | (*b*) Report category boundaries when continuous variables were categorized | N/A |  |
|  |  | (*c*) If relevant, consider translating estimates of relative risk into absolute risk for a meaningful time period |  |  |

Continued on next page

| Other analyses | 17 | Report other analyses done—eg analyses of subgroups and interactions, and sensitivity analyses 8, We conducted sensitivity analyses using a higher threshold of at least two microhaemorrhages at any MRI visit (Supplementary File 1). |  |  |
| --- | --- | --- | --- | --- |
| Discussion | | | | |
| Key results | 18 | Summarise key results with reference to study objectives |  | 6-9 |
| Limitations | 19 | Discuss limitations of the study, taking into account sources of potential bias or imprecision. Discuss both direction and magnitude of any potential bias | 8 |  |
| Interpretation | 20 | Give a cautious overall interpretation of results considering objectives, limitations, multiplicity of analyses, results from similar studies, and other relevant evidence |  | 6-9 |
| Generalisability | 21 | Discuss the generalisability (external validity) of the study results |  | 6-9 |
| Other information | |  | | |
| Funding | 22 | Give the source of funding and the role of the funders for the present study and, if applicable, for the original study on which the present article is based |  | 12 The funders had no role in the design, analysis, interpretation, or reporting of the present secondary analysis. |

*Give information separately for cases and controls in case-control studies and, if applicable, for exposed and unexposed groups in cohort and cross-sectional studies.

**Note:** An Explanation and Elaboration article discusses each checklist item and gives methodological background and published examples of transparent reporting. The STROBE checklist is best used in conjunction with this article (freely available on the Web sites of PLoS Medicine at http://www.plosmedicine.org/, Annals of Internal Medicine at http://www.annals.org/, and Epidemiology at http://www.epidem.com/). Information on the STROBE Initiative is available at www.strobe-statement.org.
