## Supplementary figures and images for "Location-specific Longitudinal Patterns of Subcortical and Lobar Cerebral Microhemorrhages in Preclinical Alzheimer Disease"

### Supplementary File 2

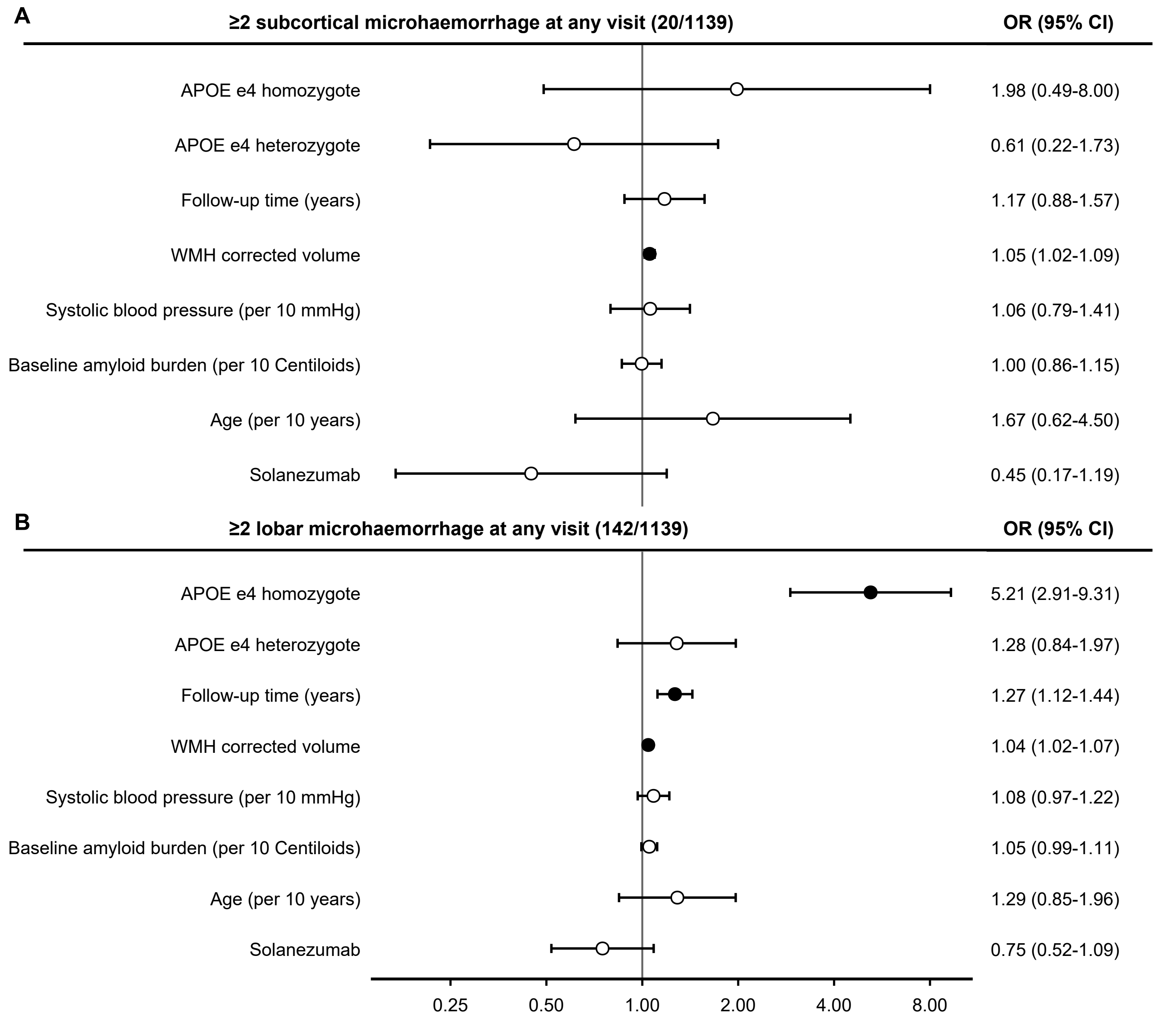
